# Perception of emergent epidemic of COVID-2019 / SARS CoV-2 on the Polish Internet

**DOI:** 10.1101/2020.03.29.20046789

**Authors:** Andrzej Jarynowski, Monika Wójta-Kempa, Vitaly Belik

## Abstract

**Problem:** Due to the spread of SARS CoV-2 virus infection and COVID-2019 disease, there is an urgent need to analyze COVID-2019 epidemic perception in Poland. This would enable authorities for preparation of specific actions minimizing public health and economic risks.

**Methods:** We study the perception of COVID-2019 epidemic in Polish society using quantitative analysis of its digital footprints on the Internet (on *Twitter, Google, YouTube, Wikipedia* and electronic media represented by *Event Registry*) from January 2020 to 12.03.2020 (before and after official introduction to Poland on 04.03.2020). To this end we utilize data mining, social network analysis, natural language processing techniques. Each examined internet platform was analyzed for representativeness and composition of the target group.

**Results:** We identified three temporal major cluster of the interest before disease introduction on the topic COVID-2019: China- and Italy-related peaks on all platforms, as well as a peak on social media related to the recent special law on combating COVID-2019. Besides, there was a peak in interest on the day of officially confirmed introduction as well as an exponential increase of interest when the Polish government “declared war against disease” with a massive mitigation program. From sociolingistic perspective, we found that concepts and issues of threat, fear and prevention prevailed before introduction. After introduction, practical concepts about disease and epidemic dominate. We have found out that Twitter reflected the structural division of the Polish political sphere. We were able to identify clear communities of governing party, mainstream oppostition and a protestant group and potential sources of disinformation. We have also detected bluring boundaries between comminities after disease introduction.

**Conclusions:** Traditional and social media do not only reflect reality, but also create it. Due to filter “bubbles” observed on Twitter, public information campaigns might have less impact on society than expected. For greater penetration, it might be necessary to diversify information channels to reach as many people as possible which might already be happening. Moreover, it might be necessary to prevent the spread of disinformation, which is now possible in Poland due to the special law on combating COVID-2019.

## INTRODUCTION

Although a large part of the Polish population has heard about coronaviruses for the first time a few weeks ago, in reality they face less dangerous coronaviruses causing simple cold all the time. Only the emergence of a novel strain from Wuhan gave the word “Coronavirus” a new meaning. Before disease introduction, less than a half of surveyed Poles believed that corona virus is the most important topic in the second half of February 2020 (IBRIS [1]). The disease was not detected in Poland until 03.03.2020, but this topic was relatively important already before introduction and started to drive media and social life after the disease introduction. During opinion poll performed on 09.03.2020 and 10.03.2020, 63% of respondents reported, that this is a serious threat for Poland, 40% that this is a serious threat for their family and 73% that this is a serious threat for the Polish Economy ([2]). We observe several small peaks of interest on the Internet for Coronavirus in Poland during investigated period (since January 2020 till 11.03.2020), although the largest surge in interest occurs after the official confirmation of the virus introduction to Poland on 04.03.2020. The last day of analysis 11.03.2020 was a day of pandemic declaration by WHO.

Recently social media activities are being analyzed worldwide to better understand perception and spread of diseases. This helps in some cases to track the spread of diseases (Ginsberg et al. [3], Lu et al. [4], Joshi et al. [5]) with a higher precision than other methods. The Internet is a good ground for propagation of views often contradicting the current state of medical knowledge [6]. Social media can serve as a valuable source of information as well as disinformation about the virus globally, fueling panic and creating so-called infodemic ([7]), at unprecedented speed and massively desrupting entire countries such as Italy (Guardian [8]). Propaganda and persuasion techniques are widely used on the Internet easily reaching certain target groups susceptible to conspiracy theories and effectively polarizing societies, possibly due to the interference by foreign intelligence [9]. Panic-related behaviors accompanying a virus outbreak [10] are influencing the epidemic spread (Perra et al. [11], Fenichel et al. [12], Wang et al. [13]), with the Internet being the main mediating mechanism. This could lead to destructive collective behavior, such as e.g. xenophobia against people from affected countries.

### Methodology

In this study, by quantitative analysis of digital traces on the Internet (e.g. social media), we concetrate on the number and nature of social media events such as information queries and deploy social network analysis, as well simple natural language processing topic modeling techniques.

Up to our knowledge there were no previous studies quantitatively linking the Internet activities and risk perception of infectious diseases in Poland ([14, 15]), expect our own study ([16]). Thus the present study is a first exploratory attempt filling this gap to continuing prelimiary research on Coronovirus perception in Poland before disease introduction ([16]).

We primarily analyze quantitative digital footprint data on the Internet from January 2020 to 11.03.2020, including their representativeness. The number of internet users in Poland in January 2020 was 28.1 million (PBI [17]) and 28.6 milion in 2019 (PBI [18]). Internet covers 85% of the total literate population in Poland. Thus, the passive representativeness of the Internet is relatively high, but active (own content creation) is biased towards younger age groups and women. The former group has a high activity on the Internet, e.g. an average Polish teenager spends about 5 hours a day on the Internet (Tanasś et al. [19])) and the later group is responsible for generation of up to 85% of health-related content in social media((PBI [17]). Over 99% of young Polish women use the Internet (Jarynowski and Belik [20])).

As a keyword in queries of web services ([15]) we chose a colloqial term “Koronawirus”/”Coronavius” due to its penetration in the society. Other related keywords in use are much less popular, except for Wikipedia, where medical term SARS-CoV-2 was choosen.

In our study each considered internet platform is described separately and has its own specific bias. Data can be biased e.g. due to content presenting algorithms on the media platforms. For instance, technological giants Google, Twitter and Facebook are supposed to implement fact verification algorithms to filter out false informations. Being aware of this, computational techniques of social sciences (Jarynowski et al. [21]), despite some disadvantages and their exploratory nature, provide us with an opportunity to analyze a huge amount of digital footprint data at low cost and in short time.

## GOOGLE TRENDS

Google has 95% share among Polish Internet users with over 8 billion entries per month and is the undisputed leader on the Polish Internet market (PBI [17]). Interest in novel Coronavirus on Google can be measured by the number of queries [Fig. 1]. According to Google there were 2 · 10^5^ searches of term “Koronawirus” monthly. However, it is calculated based on historical data. Thus there were dozens of thousand daily searches in the late February 2020.

**Figure 1.**
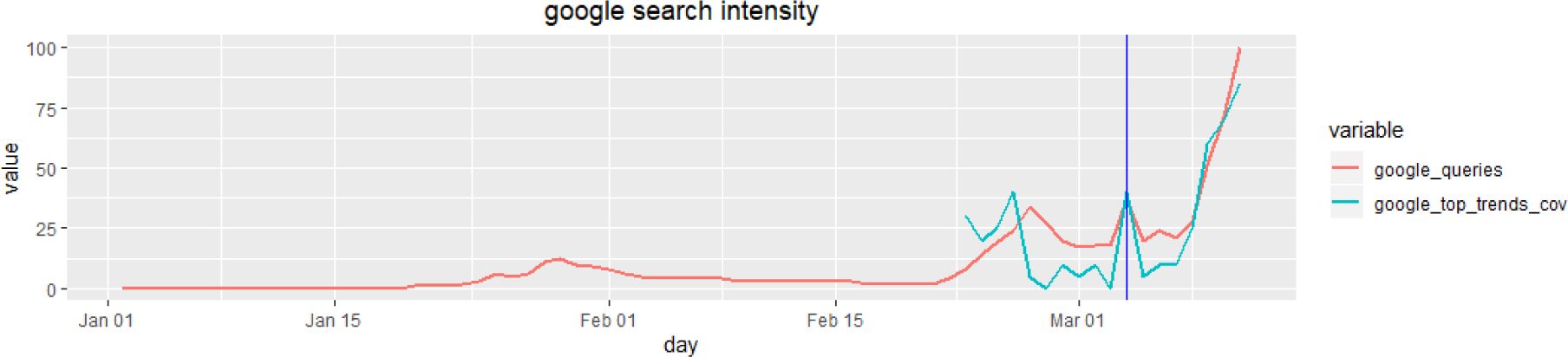
The intensity of searched queries with the word “corona virus”/”Koronawirus” in Polish Google (01.01-11.03.20) and percentage of “coronavius” related queries in top trend topics (23.01-11.03.20) both generated using Google Trend tool. Disease introduction marked with the blue vertical line.

The subject of the corona virus occurred recently after the outbreak in Wuhan. It is important to note, that until disease introduction to Poland there were no Coronavirus related searches in top 25 Google queries at all. Although “coronavirus” related queries were observed in top trends before the introduction of the disease to Poland, they start to dominate top queries only after the massive mitigation measures were taken such as school/university and boarder closures around 09-12.03.20 [Fig. 1]. Prior to the disease introduction two phases of interest can be distinguished [Fig. 1]:

- from the end of January and beginning of February when the epidemic was announced and confirmed in China. We see a small peak around 25.01.20 (e.g. death of Liang Wudong) and around 29.01.20 (e.g. first case in Germany);
- from the end of February till beginning of March (when the number of infections rapidly increased in Italy). We see a clear peak around 27.02.20 (e.g. fake news about possible introduction of the disease to Poland [22]).

After the first confirmed case in the country we can see a peak on that day (04.03.20) and substantial growth after important measures were implemented by Polish authorities (09-11.03).

People are looking for information on further epidemiological topics related to infection and epidemics as well [Fig. 2]. It should be noted that professional vocabulary such as “hand hygiene” practically does not appear in queries (below the “noise threshold” compared to other epidemiological terms [Fig. 2]). Queries related to hand hygiene had a peak in late February and beginning of March. Queries related to masks had their peak of popularity at the end of February and the lack of increase in popularity in March (compared to other epidemiological queries [Fig. 2]) may be due to the success of information campaigns on their alleged low effectiveness or simply due to the lack of the masks on the market.

**Figure 2.**
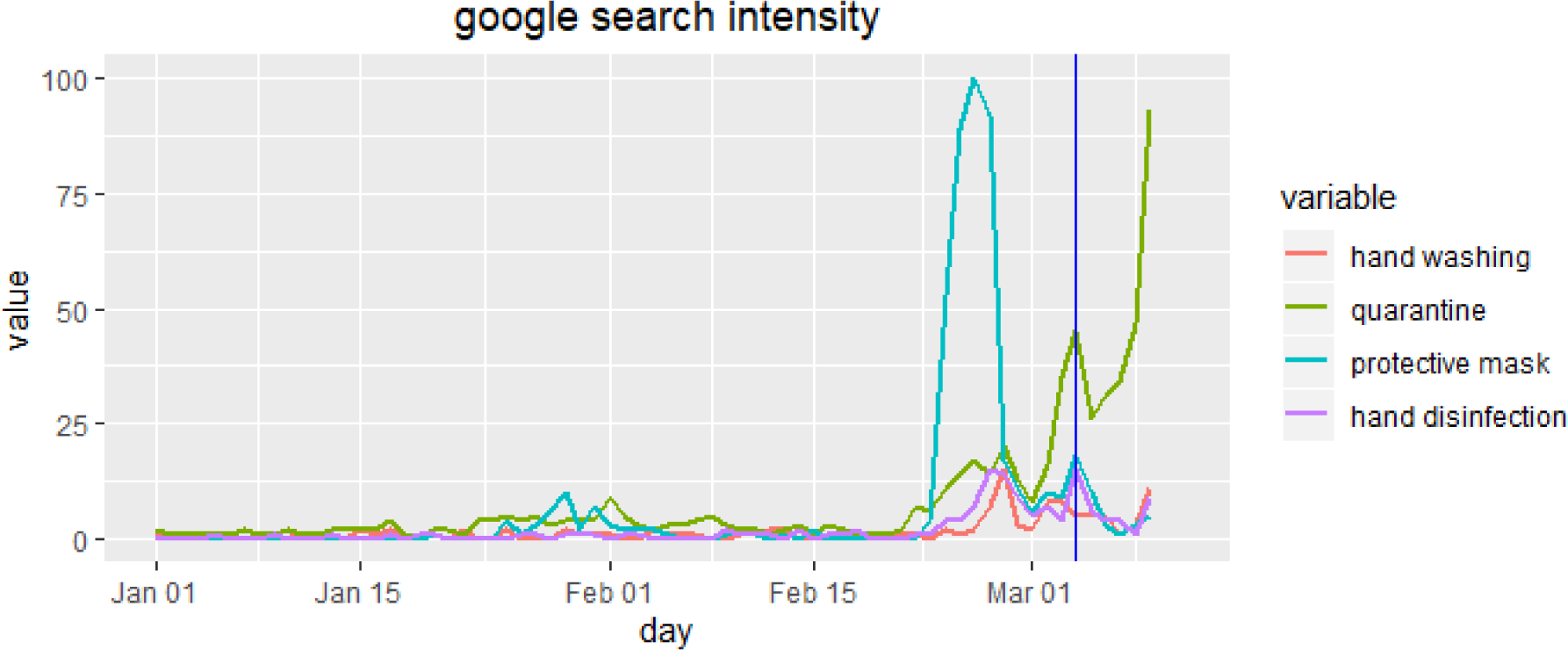
The intensity of queries with the phrases “quarantine”, “protective mask”, “hand washing”, “hand disinfection” (“kwarantanna” “maseczka ochronna”, “mycie raąk”, “dezynfekcja raąk”) in Polish Google (01.01-10.03/2020) generated using the Google Trend tool. Disease introduction marked with the blue vertical line.

Search activity for information on the infection in Poland is still much lower than in countries with high global mobility (Lai et al. [23]) and already confirmed cases of infection. Such a peripheral location of Poland (among others) lead to the first confirmed case introduced by land rather than by air (Interdisciplinary [24]).

In the corresponding semantic networks we recognize the most common co-occurring phrases in the search engine together with the word “Coronavirus” [Fig. 3]. Such a network contains information on how the predictor - the corona virus noun - relates to its arguments in a query phrase ([25]).

**Figure 3.**
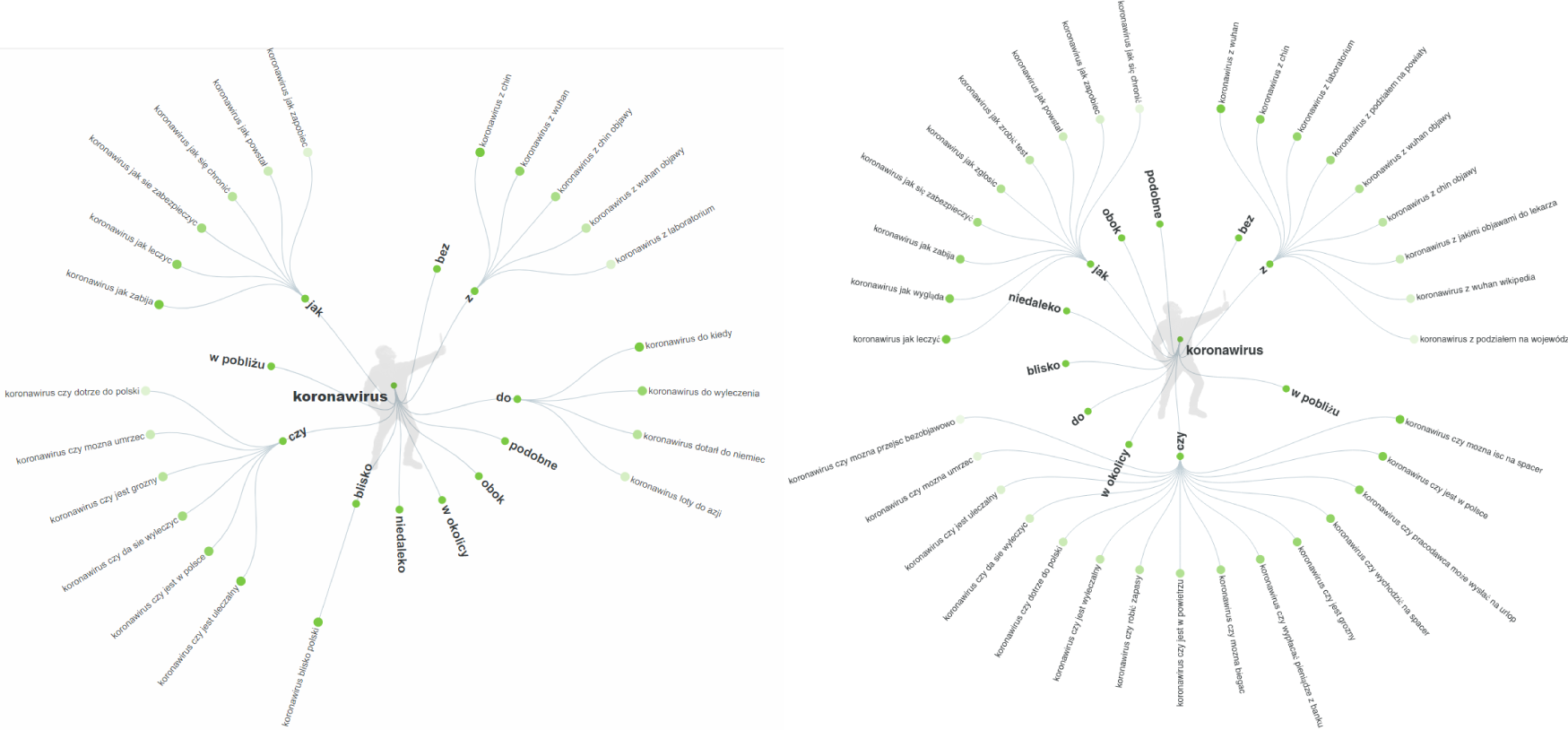
Semantic web (prepositions) of the noun “corona virus” in Google search engine before introduction (snapshot on 29.02.20) [left] and after introduction (snapshot on 12.03.20) [right] generated using the Answer the Public tool for Poland and Polish language (AnswearPublic [26]).

Before virus introduction, we observed [Fig. 3], that most often the questions are associated with a threat, e.g. if *it is / will reach / to Poland / close to Poland; can one die / how it kills* (czy jest / dotrze w / do Polski(ce) / blisko Polski; czy mozżna umrzecć / jak zabija). Second level searches concerns prevention, e.g. *how to prevent / guard / protect yourself* (jak zapobiec / chronicć si ę / zabezpieczycć si ę). In addition, there are third level threads such as symptoms, history, or restrictions. The aspect of geographical proximity is also very important, thus the terms *nearby, near* next to dominant semantic field around the word “Coronavirus”.

After virus introduction, there is domination of practical questions e.g. *how to do the test / when to call a doctor / how to treat / asymptomatic course / can you go for a walk / is it in air/ do you do shopping / leave from the employer / map* (jak zrobicć test/ kiedy dzwonicć po lekarza/ jak si ę leczycć/ przebieg bezobjawowy/ czy mozżna isćć na spacer/ czy jest w powietrzu/ czy robicć zasapy/ urlop od pracodawcy/ mapy).

## WIKIPEDIA

Wikipedia traffic is another indicator of the social activity. Wikipedia has an Internet coverage of 57% with over 350 million entries per month among Polish Internet users (PBI [17]). There is a significant overepresentation of users with tertiary education inhabiting big cities (affinity index>120 ([27])). We looked at the history of page views on discussions around the articles “SARS-CoV-2” ([28]) and “Spread of virus infection SARS-CoV-2”/”Szerzeniesi ężakazżenń wirusem SARS-CoV-2” (on 11.03.2020 this page was taken down and new page about the pandemic appeared in its place ([29])) in the Polish Wikipedia [Fig. 4].

**Figure 4.**
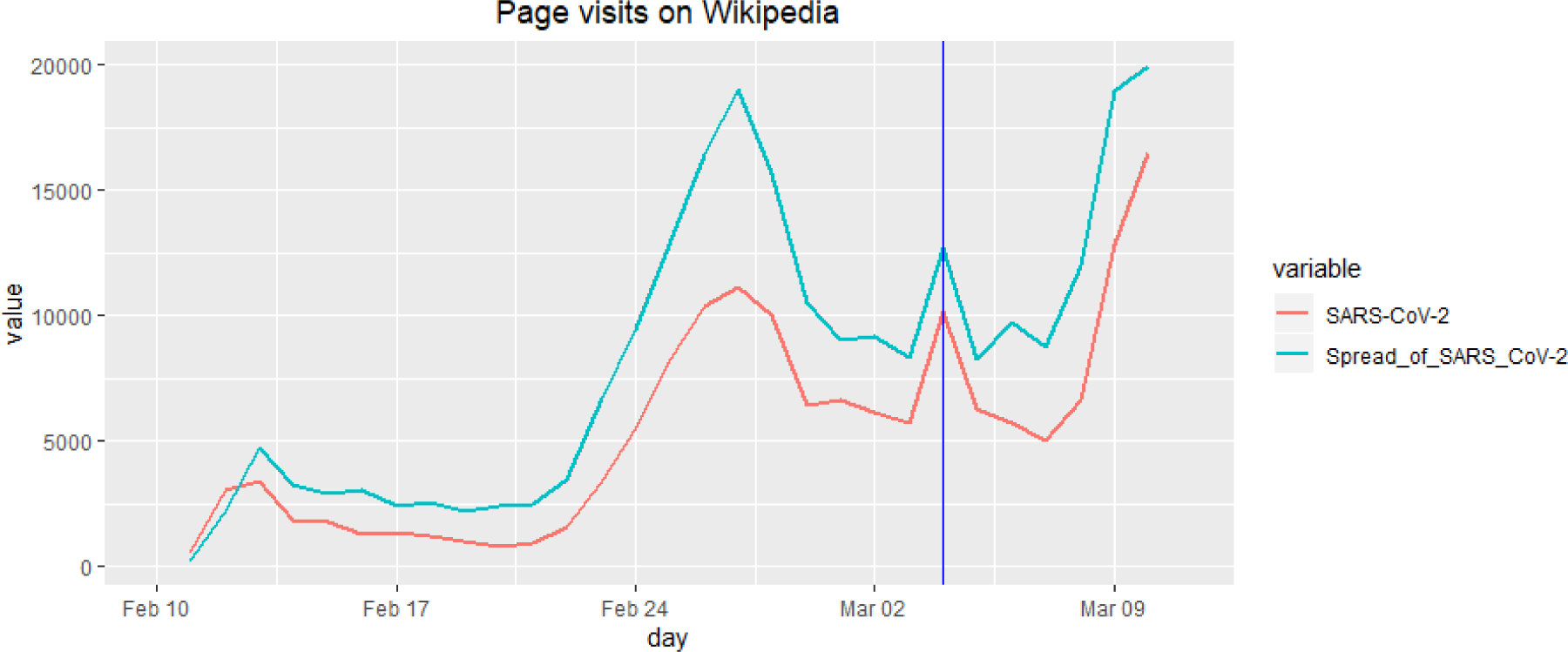
The number of views on the article “Spread of SARS-CoV-2 virus infection”/ “Szerzenie si ę zakazżenń wirusem SARS-CoV-2” and “SARS-Cov-2” (10.02-10.03.2020) on Wikipedia. Disease introduction marked with the blue vertical line.

Before the disease introduction, we see a growing trend in the number of queries with a small peak around 13.02.20 (which does not appear in other media and is related probably to giving a new name to the virus and the disease) and a clear peak around 27.02.20 (e.g. a fake news about disease in Poland ([22])). The first days of March are characterized by a slight decline in interest, perhaps due to the saturation of knowledge of basic definitions about the disease in the society. After the first confirmed case, there is a small peak around this day and a slight growth during actual epidemic and massive mitigation strategies. On 04.03.20, a new page for epidemic spread in Poland was launched.

The intensive discussions between editors in Wikipedia concern, among other, the effectiveness of protective masks or the reliability of data from the PRC (People’s Republic of China). No data is available before 10.02.2020, due to changes in the title of the articles together with the name of the virus and disease by WHO.

## EVENT REGISTRY

We choose EventRegistry (EventRegistry [30]) as a traditional media search engine because it has a large range of online magazines representing various political sites. In addition, it gives priority to the digital versions of other broadcasting channels, including television, radio or newspapers. Between January 31 and March 11, 10755 (before 4603 and 6152 after introduction) representative articles were selected (the non-systematic sampling method was applied).

In the traditional media the weekly seasonality of the articles and 3 peaks of interest: at the end of January, the second half of February and the beginning of March can be cleary seen [Fig. 5]. Counts of news article seem to coindence with all peaks observed at other platforms and expect the peak on 26.02, ahead all of other platforms.

**Figure 5.**
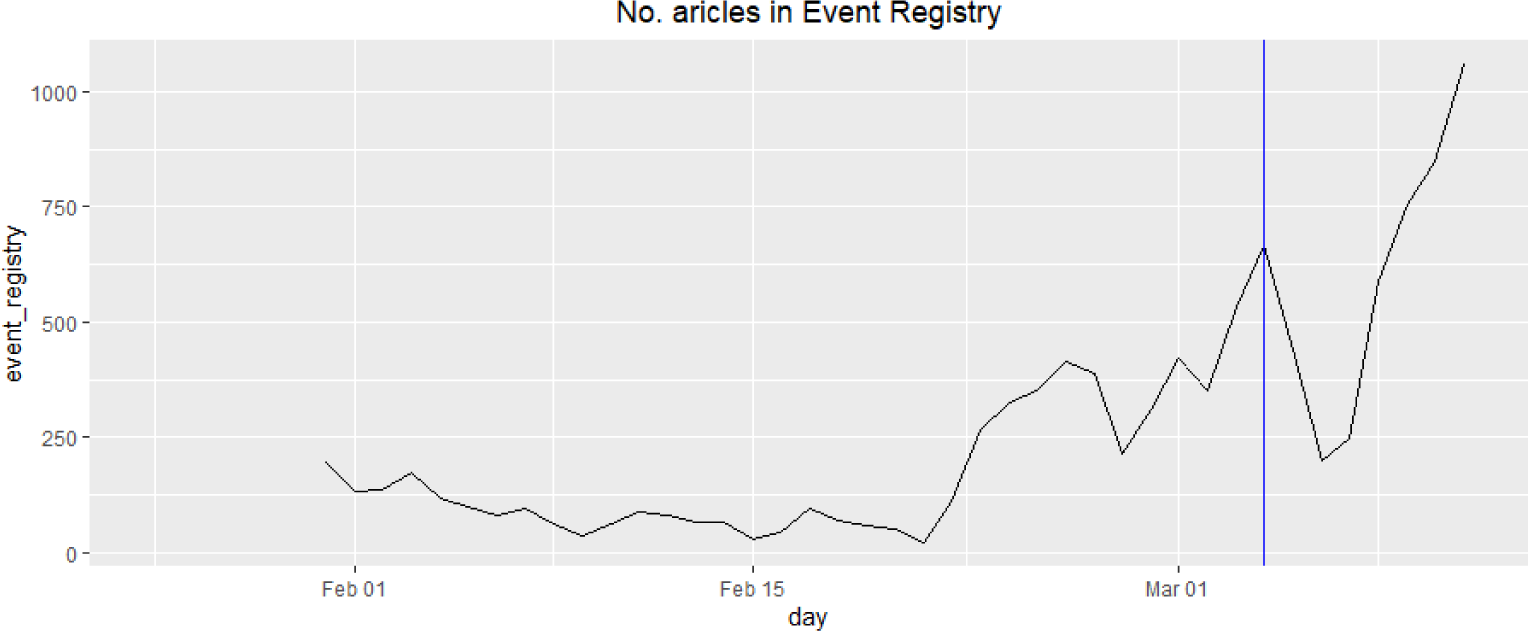
Number of articles in time 31.01-11.03 (generated using the Event Registry tool). Disease introduction marked with the blue vertical line.

## TWITTER

Twitter in Poland has relatively low popularity (∼3 million registered users or less than 8% of the population) and is mainly used by expats, journalists and politicians (Sotrender [31]). However, Twitter provides an API for data acquisition available to general public almost for free. This makes possible to analyze not only content of tweets, but also their context (location, retweeting, commenting, etc.). A lot of interest in infection can be seen on Twitter in Polish (210182 tweets with #Koronawirus during 28.01-11.03.2020).

There is not so much attention on Twitter until late February 2020. There are peaks on 27.02.20 (fake news about possible case in Poland), 02.03.20 (discussion about special anti-COVID-2010 act ([32])), 04.03.20 (disease introduction) and huge increase in interest on 9-11.03.20 (massive mitigation strategies implemented).

To apply Social Network Analysis (Wasserman et al. [33]) methods to the Twitter data, we build a network with vertices representing Twitter accounts and edges representing retweets (Jarynowski et al. [34]). The network revealed various connections (social impact, trust, friendship, etc.) between accounts being social actors and the characteristics of the actors (political affiliation, views, etc.). An unsupervised weighted Louvain algorithm (Blondel et al. [35]) for community analysis was used and the vertex color denotes the community it belongs to. Retweet network [Fig. 7] shows how the discourse is divided into the ruling party (gray), opposition (orange) and the protestant religious and political group / "Idz pod prad” (light blue).

**Figure 6.**
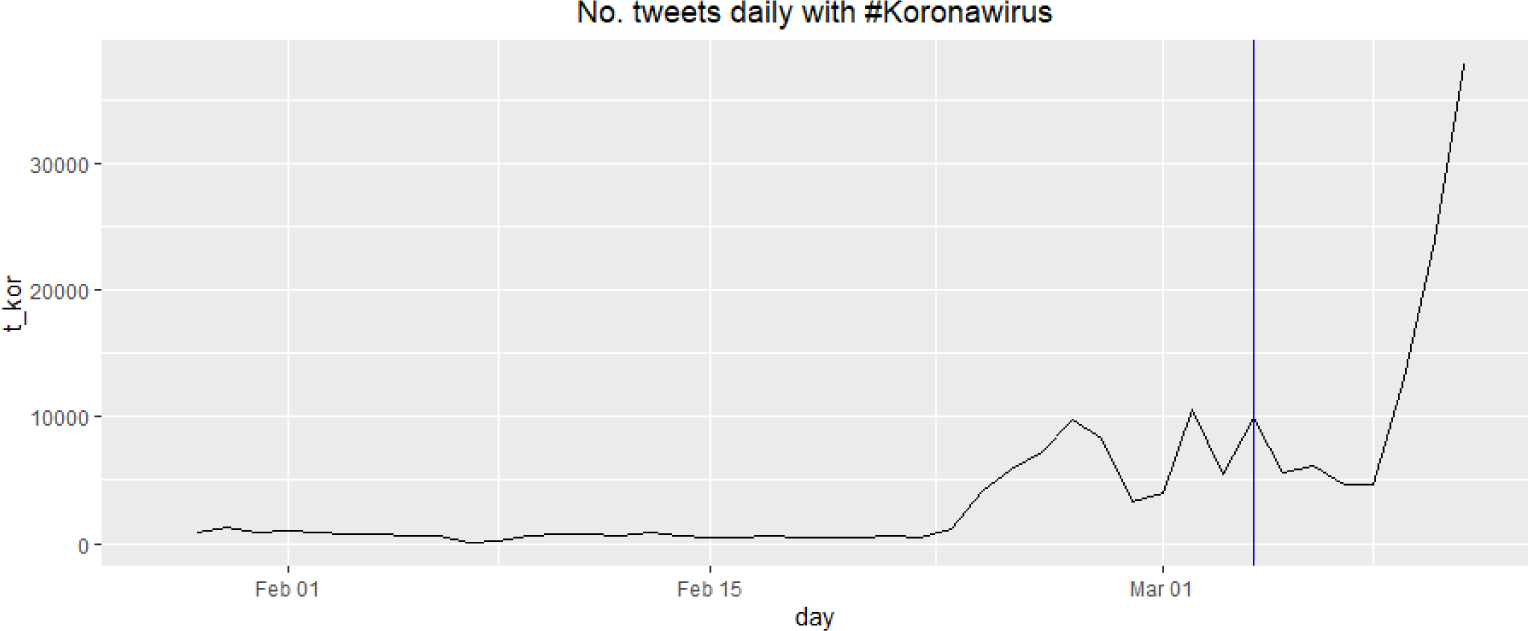
Number of tweets per day with the Koronawirus hashtag in Polish language (28.01-11.03.2020). Disease introduction marked with the blue vertical line

**Figure 7.**
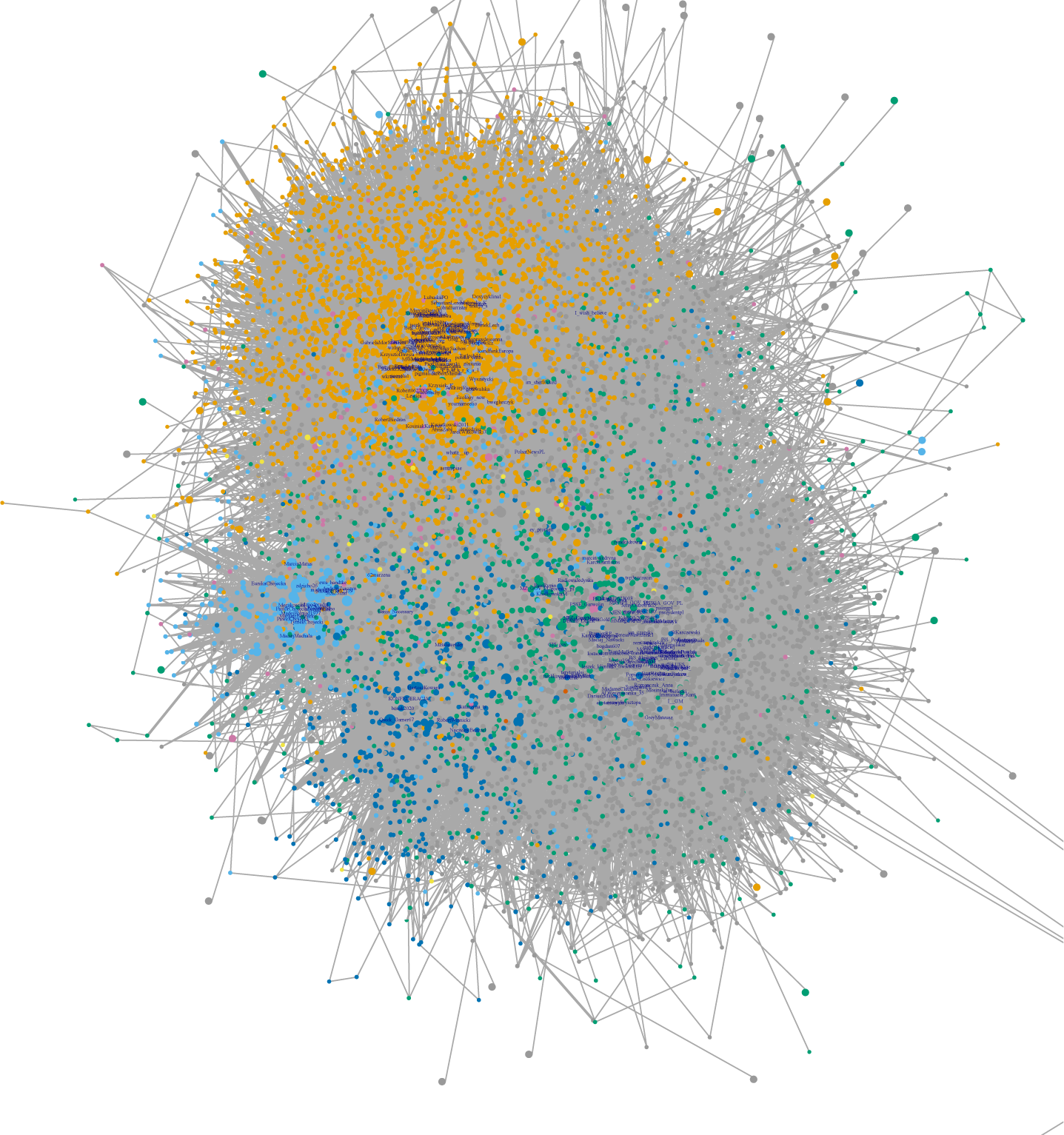
A network of Twitter accounts (vertices) connected by retweets (edges) with the Koronawirus hashtag. Colorcode: gray - the ruling camp, orange - the opposition, light blue - the Protestant group, dark blue - far right (28.01-11.03.2020). This network only shows accounts that have generated at least 3 tweets and connections that represent at least 2 retweets, labels are provided fo 100 most central (weighted degree centrality).

In addition, further discourse participants were identified [Fig. 7] as far right (dark blue). The subject of “Coronavirus” in Poland has a conflict-generating potential. As a consequence a dispute has emerged between the ruling party promoting information content and confirming the belief that the Polish state is well prepared to fight the virus (gray cluster), and the opposition negating its ability to fight the virus [Fig. 7], especially before disease introduction (Jarynowski et al. [16]). However after introduction, the most distinguished discourse is represented in the cores of the network [Fig. 7] and boundaries between communities are more blured than it was before confirmation of the first case in Poland ([16]). It could mean, that (at least in this weekly resolution) for an average Twitter user, mechanisms of solidarity and community building (for example to help neigbors and support healthworkers) are less politically driven than before.

For example, Twitter accounts already classified as potentially belonging to the so-called trolls (which in other studies were attributed to the extreme right in the context of elections to the European Parlament (OKO [36]), or to the far-left side in the context of the African Swine Fever epidemic (Jarynowski et al. [37]), promoted content in the buffer area (attacking both the ruling party and the mainstream opposition).

We also looked on the most frequent words after stemming and excluding stop words [Tab. I]. There are mainly words related to topics around health, but with politicical context as well, due to the political and journalistic bias of Twitter. In comparition to situation before the first confrimed case ([16]), there is less politics and less fear related concepts.

**Table I.** Counts of 30 most frequent words on Tweeter related to COVID-10 without stop words during 28.01-11.03.2020.

| Rank | item | count | item English | Rank | item | count | item English |
| --- | --- | --- | --- | --- | --- | --- | --- |
| 1 | polska | 32266 | Poland | 16 | informacja | 12248 | information |
| 2 | rzad | 22500 | Government | 17 | szpital | 12217 | hospital |
| 3 | przypadek | 21868 | case | 18 | szkola | 12168 | school |
| 4 | osoba | 18723 | person | 19 | chiny | 12153 | China |
| 5 | polski | 18482 | Polish | 20 | zarazic | 12127 | to infect |
| 6 | wszystek | 17924 | all | 21 | epidemia | 12117 | epidemic |
| 7 | móc | 17908 | can | 22 | wirus | 11349 | virus |
| 8 | zdrowie | 17476 | health | 23 | walka | 10904 | fight |
| 9 | wlochy | 14113 | Italy | 24 | zakazic | 10888 | to infect |
| 10 | czlowiek | 13681 | men | 25 | kwarantanna | 10552 | quarantine |
| 11 | wszyscy | 13586 | all | 26 | potwierdzic | 10423 | to confirm |
| 12 | minister | 13079 | Minister | 27 | dzien | 10393 | day |
| 13 | test | 13079 | test | 28 | polak | 10341 | Pole |
| 14 | pis | 12810 | Law and Justice | 29 | panstwo | 9769 | country |
| 15 | ludzie | 12602 | people | 30 | zamknac | 9713 | to close |

## YOUTUBE

Youtube has a 68% share among Polish Internet users with about 700 million entries per month (PBI [17]). In addition, streams from the mobile app should be taken into account, as it is most popular app on Poles’ smartphones (PBI [17]). For our analysis, we selected videos on the main subject of “corona virus” ([38]) using keyword search. One observes multiple peaks (25.01, 31.01, 27.02, 2.03, 04.03, and 9-11.03.20) on Youtube similar to all other social media [Fig. 8].

**Figure 8.**
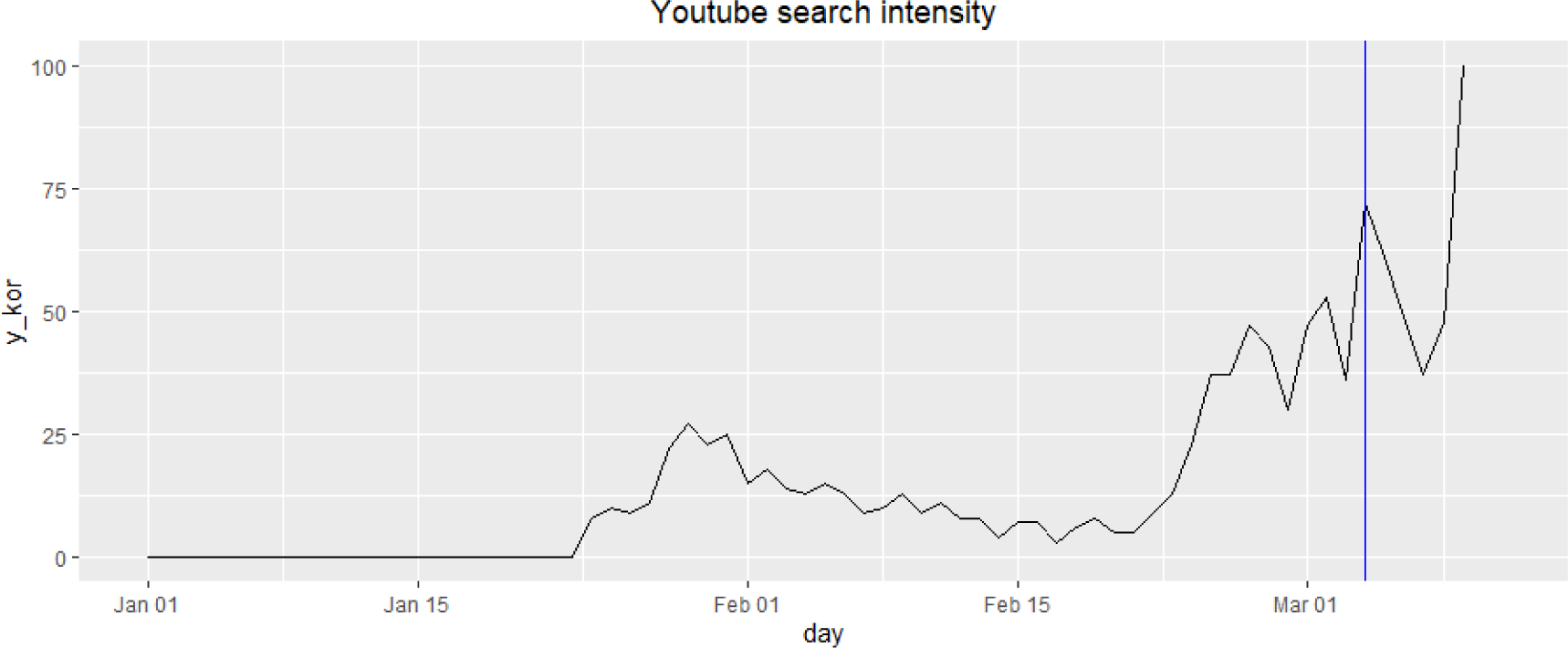
The intensity of queries for the word “Koronawirus” on Youtube (01.01-11.03.20) generated using the Google Trend tool. Confirmed disease introduction is marked by the blue vertical line.

## OTHER MEDIA

The fastest growing profile in January and February 2020 in the entire Polish Facebook was “Conflicts and global disasters” ("Konflikty i katastrofy sświatowe”) which gained over 120 thousand followers in one month correlating with recent increased activity related to corona virus information. The most popular post in the Vlog category in January 2020 was a video material titled “Wuhan market” on SA Ward ęga’s profile (@sawardega), which was eventually marked as containing false information (Sotrender [39]).

There are other social platforms not covered by this study such as Instagram (a popular platform among teenagers with affinity index>120 in this age category ([40])). The topic of Coronavius was not so popular at Polish Instagram until the disease introduction. The most popular hashtag after introduction of the virus “odwolajcieszkolyxd” and is related to school closures.

For a better topic coverage blogs (e.g. blog Media@jesion gathers hundreds thousands entries by day ([41])) and user comments in Internet media presents additional reach information source. E.g. articles from media such as wp.pl, onet.pl, interia.pl have billions entries monthly and their articles on Coronavirus reaches on average dozens of thousands entries with hundreds of comments ([17]).

## COMPARISON OF DIFFERENT PLATFORMS

To compare the interest on COVID-19 on different Internet platforms we visualized the available queries and interest measurements as time series together and marked events important to the Polish public [Fig. 10]. From the time series as well as Google queries semantics [Fig. 3] and tags/topics in the news, we observe that geographal proximity of the disease drives a lot of interest (e.g. outbreak in Germany). We see that traditional news agencies (well represented in EventRegistry) as well as Google search could anticipate and form more distinct interest peaks than social media platforms. Moreover, discussions on social-content media (YouTube and Wikipedia) cause smoother interest trajectories [Fig. 10] than information providers (news from EventRegistry, Google). We detected time lags between different platforms for given topics (e.g news from event registry are ahead of commentatory media in fake news on possible disease introduction). It could mean that the social media dissiminate infomation via word-of-mouth speading mechanisms. On the other hand, traditional media have journalists hired to search and select most interesting topics quickly ([42]). It leads to faster respond to events by news agencies (repersented by EventRegistry) than social media. A stronger lag (1 day) is observed between EventRegitry (the most ahead of) and Twitter (the most delayed) [Fig. 9].

**Figure 9.**
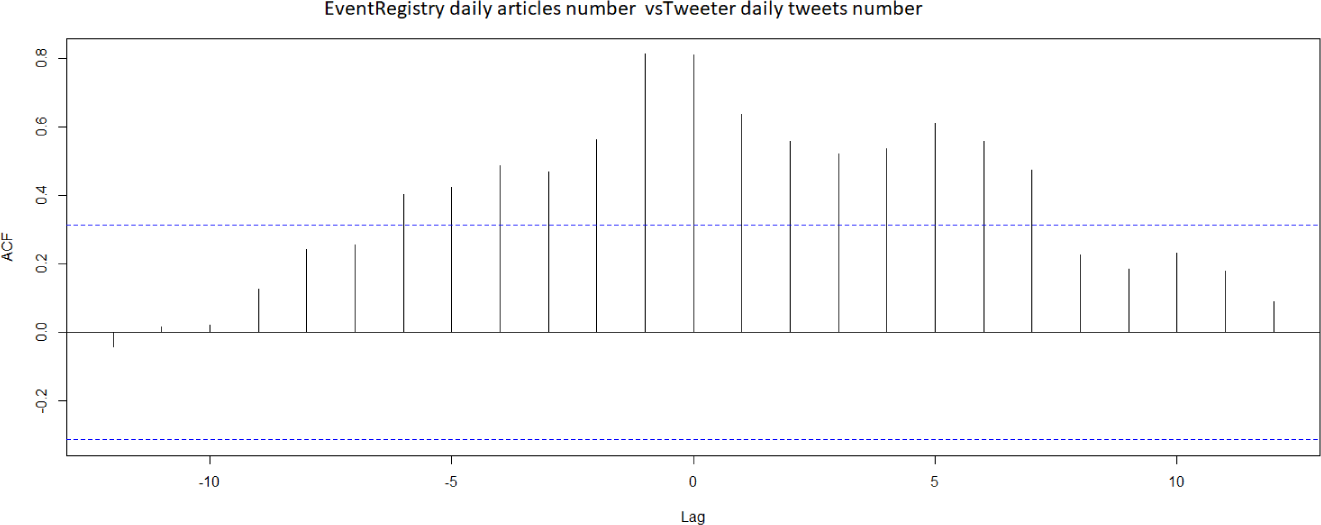
Lagged (in days) correlation between daily series of article counts from Event Registry and Tweets numbers.

**Figure 10.**
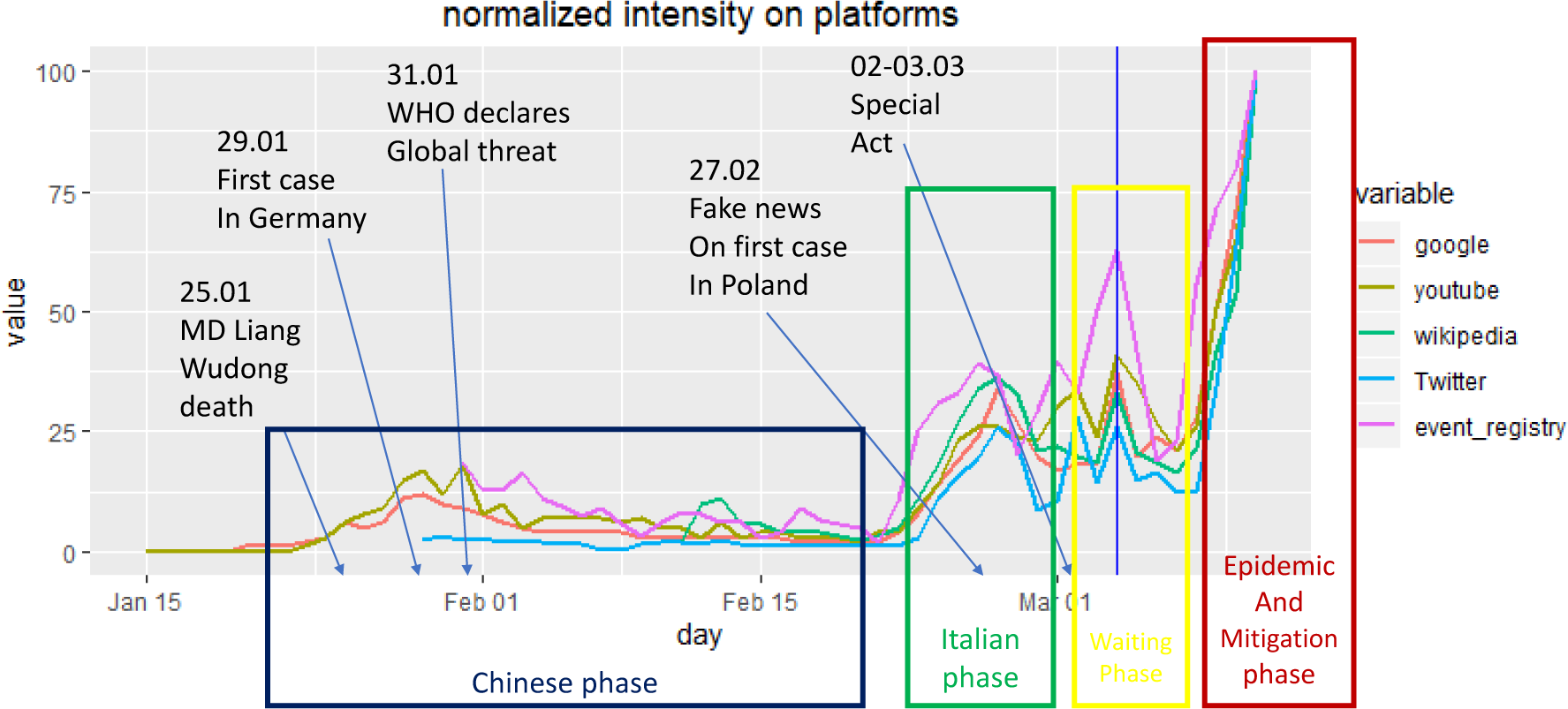
The intensity of topic Koronawirus on various media platforms during 15.01-11.03.2020. Time series were normalized to 100 by maximal value for a given serie. Disease introduction is marked by the blue vertical line.

On Fig 10 we can distinguish 4 phases of interest in Poland ([16]):

1. Chinese phase (COVID-19 emerges in Asia ([43]));
2. Itialian phase (second wave of COVID-19 ([44]));
3. Waiting phase (awearness building and waiting for a first case in Poland);
4. Epidemic and Mitigation phase ([45]).

Moreover, Twitter (15% penetration rate among Polish Internet users ([40])) has a significantly different time pattern than other analyzed platforms. In particular, it does not have China-related phase at all and it has at least 3-fold faster growth rate than other media in epidemic and mitigation phase.

To quantify the similarity in interest on different platforms we calculated Pearson correlations coefficients of the time series [Fig. 11]. All measured intensities are positively correlated. *Protective mask* as a signal of fear/perception of risk is less correlated with other variables more related to information needs. *Protective mask* and *washing hands* is much less correlated than *protective mask* and *hand disinfection*, which could mean, that people search for professianal solutions rather thans simple and practical ones. Moreover, high amounts of searching terms as *antiviral mask* (there is no such a medical term) implies that people are searching for colloquial terms ([16]).

**Figure 11.**
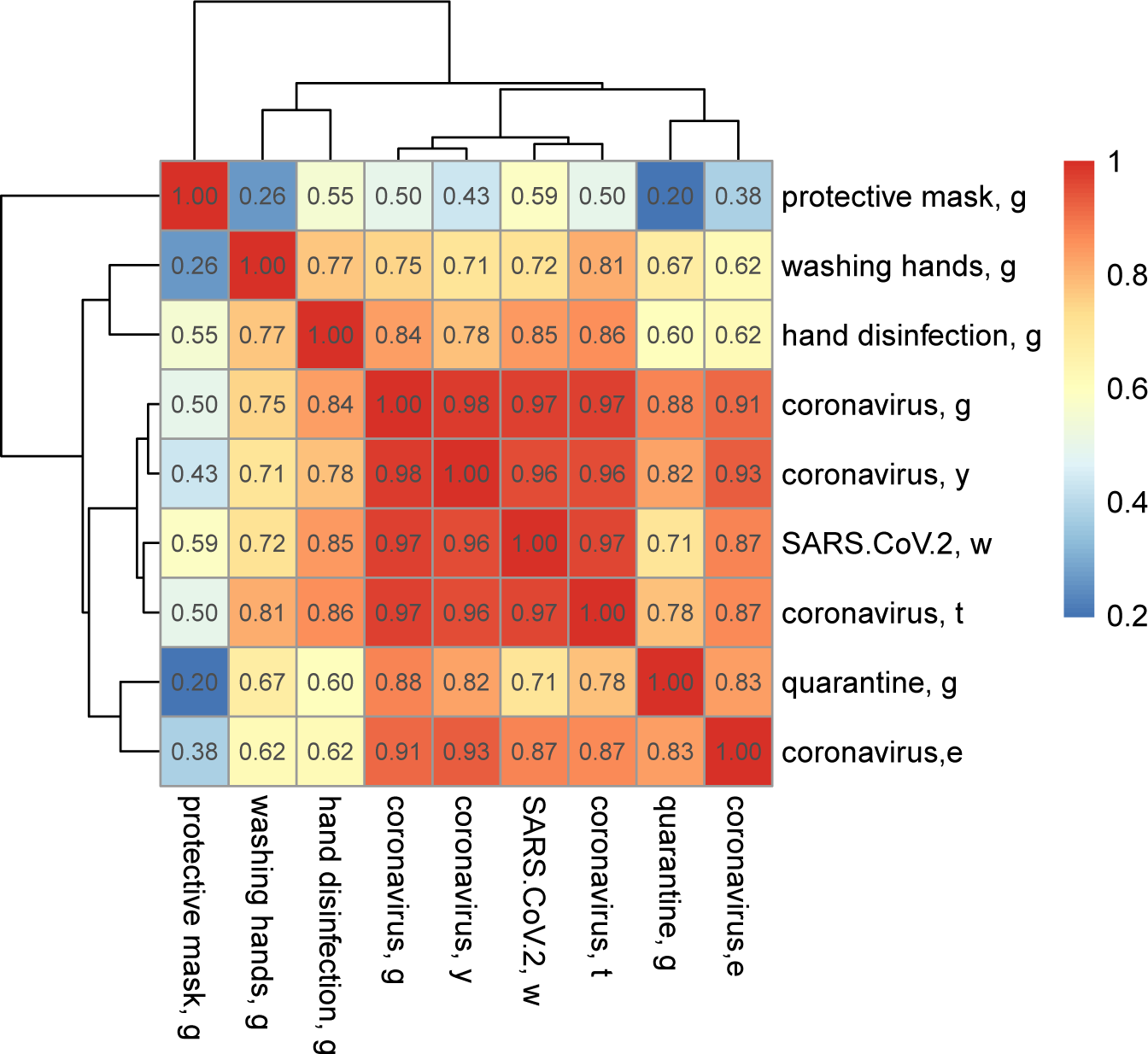
The Pearson’s correlation matrix and the corresponding hierarhical clustering for the terms “Koronawirus”/”Covronavius” and related epidemiological queries on various media platforms (g – Google, w – Wikipedia, y – Youtube, t – Twitter, e – EventRegistry). With the significance level of 5% all correlations were significant except the pair “antiviral mask, g”/”quarantine, g”. Colorcode corresponds to correlation strenght.

## CONCLUSIONS

In the face of the COVID-19 pandemic, there is an unprecedented flood of information (information ‘noise’) and our task was to extract important events and features from widest range of Internet media in Poland. The knowledge of quantitative characteristics of the “Coronavirus” perception in Poland is an important prerequisite for a proper crisis management (Trzos et al. [46]) such a design of protection policies for risk management and adequate education of citizens by the stakeholders identified in Poland. For example, at present, government mitigation programs or hygiene education principles published on Twitter fall (to the great extend) into the information bubble or echo chambers ([47]) of supporters of the ruling party [Fig. 7]. Lack of interest in general public ([43, 48]) could be associated with a low epidemiological awareness of average Poles, especially when COVID-19 pandemic is massively discussed in traditional and social media by a relatively small but loud group of people.

Before official disease introduction we observed in Poland two information phases related to outbreaks in China (the end of January) and in Italy (the second half of February) and one commentary and update phase in social media in the beginning of March related to, among others, the special act on COVID-19 mitigation [Fig. 10]. Information media (Wikipedia and Google) do not display the third phase, because probably the awareness about the virus and the knowledge on the disease has already saturated, and people are interested mainly in the current update (new infomation on COVID-19) on Twitter, Youtube, or other electronic media.

After the disease introduction to Poland, there is a peak on the introduction day and fast growth of interest in epidemic and mitigation phase [Fig. 10]. Under conditions of a market-consumer society, private goals of individuals come into conflict with responsibility of the whole society or community. For example, economic consequence of supply and demand lead to price increases of medical devices ([49]).

Due to the data availability, only by analyzing Twitter we have the full control over the methodology and research techniques (Alshaabi et al. [50]). The most difficult is to access the Facebook data, despite the highest population penetration and the largest reach in Poland ([51]). Facebook does not allow automated analysis (Facebook [52]) for the public and we can only rely on manual work-intensive research of commercial companies (Sotrender [31, 39]), whose research methodology may differ from scientific standards. However, there are attempts to use available Facebook Ads campaigns in context to “Coroanavirus” [53].

In order to prepare and manage the crisis in an optimal way, a deeper perception analysis in the form of reliable quantitative and qualitative analysis is required. Especially, according to the results of empirical research, society expects institutional activities and in the event of an epidemic, it is the “state (…) that is responsible for the poor health of the population” (Taranowicz [54]). Perhaps one of the reasons the Chinese have been so successful in controlling the spread of the infection is that social media like WeChat (Wang et al. [55], Zhang et al. [56]), or Internet forums (Liu and Lu [57]) were analyzed by algorithms (Lu et al. [58], Paul and Dredze [59], Salathe et al. [60]) with a goal to mitigate the spread of COVID-Combining the behavioral changes detected via social media analysis with the detailed information on human mobility via e.g. mobile phone tracking (Schneider et al. [61], Brockmann et al. [62], Gonzalez et al. [63]), sophisticated computational models of infectious disease spread could be implemented (Belik et al. [64], Vespignani [65], Maier and Brockmann [66], Hufnagel et al. [67], Ferguson et al. [68], Prasse et al. [69]) allowing to simulate various scenarios and assess possible human and economic losses and properly distribute the available resources. Furthermore, impact of public information campaigns could be measured by internal surveys of public opinion. In addition, Internet media analysis could fill gaps in socio-medical research on collective actions during an important public health disruption event such as infectious diseases (Jarynowski and Belik [70]).

## Data Availability

Collected data and script available
https://github.com/ajarynowski/koronawirus

## Acknowledgments

we thanks PNFN (2019-21), NCN (2016/22/E/HS2/00034), and FU Berlin (FU AvH: 08166500) for partial finanacial support and Łukasz Krzowski, Daniel Płatek, Ireneusz Skawina, Andrzej Buda, and Marcus Doherr for fruitful discussions.

